# Development of a novel CRISPR/Cas13-based assay for diagnosis of *Schistosoma japonicum* infection

**DOI:** 10.1101/2022.11.11.22282198

**Authors:** Skye R. MacGregor, Donald P. McManus, Haran Sivakumaran, Juliet D. French, Catherine A. Gordon, Pengfei Cai, Remigio M. Olveda, Malcolm K. Jones, Hong You

## Abstract

Schistosomiasis is a disease that significantly impacts public health in the developing world. Effective diagnostics are urgently needed for improved control of this disease, but current diagnostic procedures lack the requisite sensitivity, portability and cost-effectiveness needed for use in resource-poor settings. We developed a novel assay for the detection of *Schistosoma japonicum* using the CRISPR mediated diagnostic platform SHERLOCK (Specific High-Sensitivity Enzymatic Reporter UnLOCKing), combining recombinase polymerase amplification (RPA) with CRISPR and CRISPR-associated RNA-guided endoribonuclease Cas13 (CRISPR-Cas13). The assay was validated using 80 faecal samples obtained from a mouse model infected with the Philippine strain of *S. japonicum*, as well as 38 clinical faecal and 37 serum samples obtained from subjects living in endemic areas for *S. japonicum* in Northern Samar, the Philippines. CRISPR-Cas13 mediated detection was determined via fluorescent readout or colorimetric readout on a lateral flow strip. Our results demonstrate that our *S. japonicum* SHERLOCK assay is specific, sensitive and user-friendly. Although the assay does not require the specialized equipment or expertise necessary for real time PCR-based detection, which is currently the most sensitive approach for the diagnosis of helminthic infections, it achieved 93-100% sensitivity compared with the qPCR, as well as 100% specificity across all the human and animal samples tested. Although further optimisation is required before field-ready implementation, CRISPR-based nucleic acid detection shows great promise as the basis of a point-of-care (POC) diagnostic tool for clinical diagnosis and surveillance of schistosomiasis with potential extension to other helminthiases.

**Author Summary:** Parasitic helminths cause devastating diseases, including schistosomiasis, afflicting 1.5 billion people worldwide and representing a significant public health and economic burden. Currently available diagnostic tools for helminth infections are neither sufficiently sensitive nor field-friendly for use in resource-poor settings where infection is most prevalent, and advanced tools are are urgently needed for rapid mapping of helminthic diseases and monitoring control efforts. For the first time, we used the *Schistosoma* bloodfluke model to successfully establish a diagnostic assay with the CRISPR-based nucleic acid detection platform SHERLOCK (Specific High-Sensitivity Enzymatic Reporter UnLOCKing) by combining recombinase polymerase amplification (RPA) and CRISPR-Cas13 detection to diagnose schistosomiasis in humans and animals. We showed that the novel CRISPR-based assay, with its low cost of application, is capable of robust detection and is field-friendly. It exhibits similar diagnostic sensitivity as qPCR-based assays, which are currently the most sensitive approach for the diagnosis of helminthic infections, but with significantly reduced requirements for trained personnel and technical expensive equipment. Our *S. japonicum* SHERLOCK assay has the potential to fulfil key recommendations of the neglected tropical diseases (NTDs) 2021-2030 roadmap and the 2022 Guideline on the Control and Elimination of Human Schistosomiasis released recently by the World Health Organization.

## Introduction

Parasitic helminths are a global scourge but mainly impact low-income communities in developing tropical/subtropical countries. Asia and sub-Saharan Africa are infection hotspots, hosting about 90% of the world’s parasitic worm infestations [1]. Parasitic worms cause severe morbidity and mortality and result in devastating chronic disease outcomes, with significant impacts on human health and economic development. Schistosomiasis is an acute and chronic parasitic disease caused by trematode blood flukes; over 250 million people are infected and 800 million are at risk of infection in 78 countries [2]. As the second most devastating of the neglected tropical diseases (NTDs), after malaria, schistosomiasis is prioritised by the World Health Organisation (WHO) for control and elimination. One key roadblock to schistosomiasis elimination, identified in the new WHO NTDs roadmap 2021-2030 [3], is the lack of a diagnostic test that provides timely and accurate results while still being accessible, inexpensive, and able to be performed by local personnel with minimal training [4]. As schistosomiasis often occurs in remote rural areas, a point-of-care (POC)-based diagnostic is particularly relevant [4]. Deployment of a sensitive POC test will confer many benefits for control efforts by significantly reducing drug costs through targeted treatment of infected subjects rather than mass drug administration (MDA) with praziquantel, thereby reducing the risk of anthelminthic-drug resistance developing and increasing treatment compliance. Moreover, a highly sensitive diagnostic test will allow for better mapping of schistosome transmission dynamics and provide more accurate disease burden estimates. As elimination programs progress in schistosomiasis-endemic areas, ultra-sensitive diagnostic procedures will be essential to accurately record the infection status of susceptible populations to prevent re-bound infections following the cessation of MDA programs [5].

Currently, diagnosis of helminthic infections (including schistosomiasis) relies almost entirely on conventional diagnostics [6-9], including stool-based Kato-Katz microscopy, the miracidial hatching technique and serological-based procedures. These diagnostic methods lack sensitivity and/or demonstrate poor replicability with variability, and are often unreliable and labour-intensive, especially when applied in areas of low parasite prevalence/intensity [10, 11]. Recently, diagnostics based on nucleic acid amplification (such as PCR-based assays and cell-free DNA detection), have achieved a much higher degree of sensitivity and specificity [12]; however the requirements for costly laboratory instrumentation, reagents, and trained personnel have significantly hampered the practicability of widespread use of these tests particularly in remote endemic areas in developing countries. Without availability of a readily accessible, sensitive and affordable diagnostic test, this leads directly to an underestimation of active schistosomiasis cases, errors in evaluation of transmission rates, and in interpreting outcomes of MDA programs [11]. A new generation of field-friendly schistosomiasis (and other helminth) diagnostics is urgently needed to improve control efforts that can eventually eliminate the spread of parasitic worm infections globally.

Over the past 5 years, a new class of CRISPR-based diagnostics has emerged that has proven especially promising in virology (including COVID-19) and cancer diagnosis [13-16]. These systems utilise the programmable CRISPR-associated endonucleases Cas12 and Cas13, and have been shown to be ultra-sensitive, rapid, specific, cost-effective and portable, without the necessity for specialized equipment and expertise [15, 17-19]. The CRISPR-based diagnostic system typically involves amplification of target sequence using isothermal amplification methods, followed by CRISPR-Cas12/13-mediated detection that combines the binding specificity of programmable crRNAs with the collateral nonspecific enzymatic cleavage activities of Cas12 and Cas13 to cleave a reporter sensor [20] that is measured via fluorescent or colorimetric readouts. Isothermal amplification methods include recombinase polymerase amplification (RPA) and loop-mediated isothermal amplification (LAMP), both of which can amplify target nucleic acids at a single temperature - usually 37°C (RPA), or 65°C (LAMP)-making the system especially suitable for field-based application. The Cas13-based SHERLOCK platform has been shown to be able to clinically diagnose viral infections (Zika and dengue virus) at concentrations as low as 1 copy/μl [19], and with single-base mismatch specificity for detection of SARS-CoV-2 in less than 1 h [21]. Recently, successful CRISPR-Cas12/13 mediated diagnostics have been developed for the detection of unicellular parasites [including *Plasmodium* [22], *Cryptosporidium parvum* [23, 24] and *Enterocytozoon hepatopenaei* [25]] and the plant cyst nematode-*Heterodera schachtii* [26]. This revolutionary technology has the potential to fulfil the unmet and urgent needs for parasite detection recommended by WHO, but has yet to be enlisted for detecting parasitic helminth infections.

Here, we describe the development of a novel diagnostic tool for schistosomiasis using the RPA-based CRISPR-Cas13 nucleic acid detection platform (SHERLOCK) that enables robust, ultrasensitive detection of the Asiatic schistosome, *Schistosoma japonicum*. Using this *Schistosoma* bloodfluke model [27], we validated the SHERLOCK assay using faecal samples collected from mice experimentally infected with cercariae (Philippines strain) of *S. japonicum*, and human faecal and serum samples obtained from a *S. japonicum-*endemic area in the Philippines. We demonstrate that our assay exhibits a similar level of diagnostic sensitivity to qPCR, which is currently the gold standard for helminth molecular diagnosis, and that SHERLOCK detection is possible using multiple POC-applicable readout options. While further optimisation will be required before the assay and the POC-readouts can be used in the field, we demonstrate here, for the first time, the untapped potential of CRISPR for the accurate diagnosis of schistosomiasis and other parasitic worm infections.

## Results

In this study, we developed a novel assay for detection of *S. japonicum* infection using the Cas13a-based SHERLOCK platform and have demonstrated the sensitivity and accuracy of the procedure using samples collected from experimentally infected mice and humans naturally infected with *S. japonicum*. The SHERLOCK assay we developed combines RPA pre-amplification with CRISPR-Cas13 mediated detection, visualised via a fluorescent or colorimetric readout (Fig 1).

**Fig 1.**
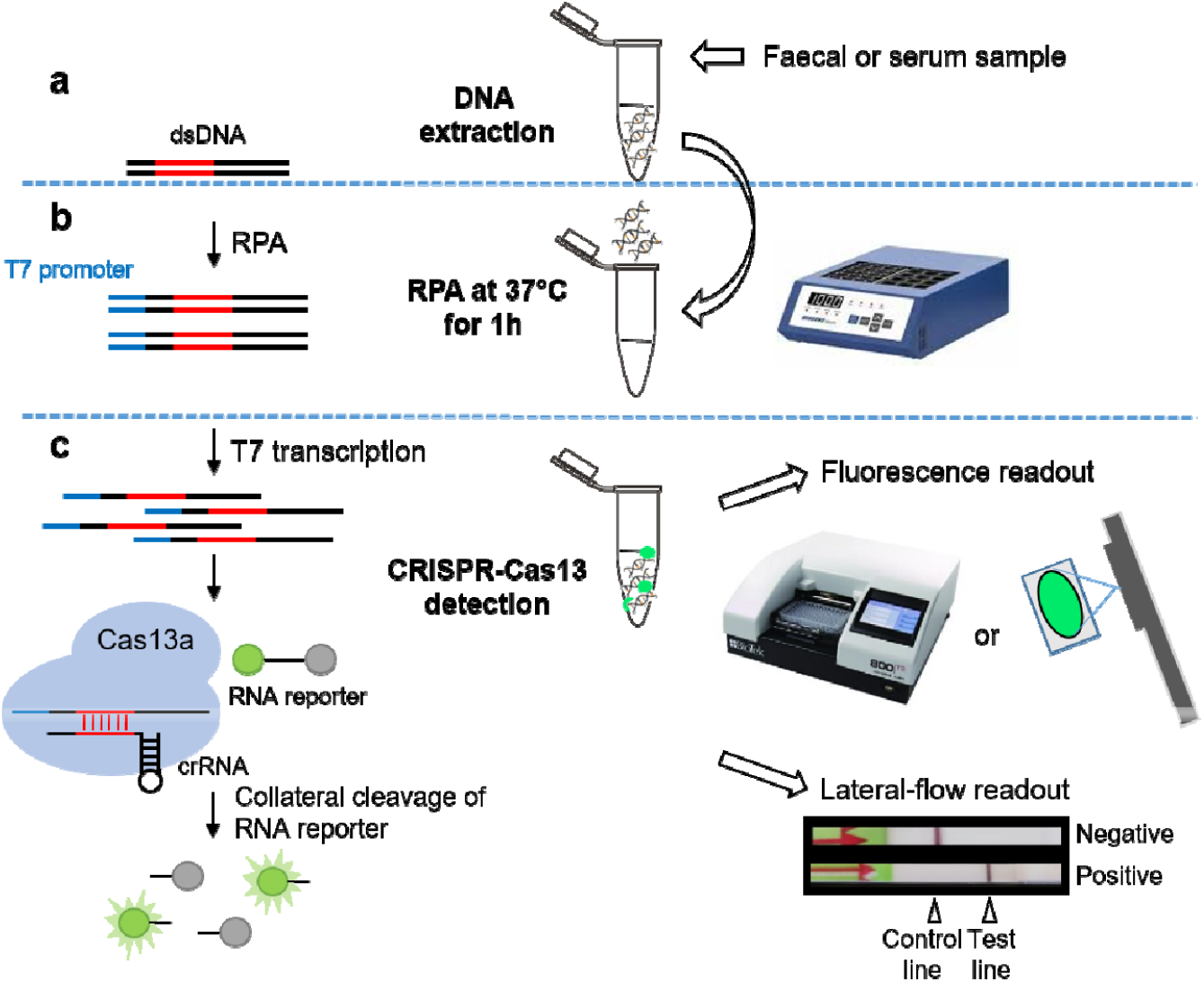
CRISPR-Cas13a SHERLOCK *S. japonicum* detection assay workflow. (a) DNA extraction from faecal or serum samples collected from human and animal; (b) Recombinase polymerase amplification (RPA) reaction performed at a single temperature (37°C) for 1 h; (c) CRISPR-Cas13a detection via either fluorescence or lateral-flow readout. The *S. japonicum* DNA target sequence is amplified by RPA. The RPA-amplified double-stranded DNA (dsDNA) is transcribed to single-stranded RNA (ssRNA) via T7 transcription. The Cas13a-crRNA complex is activated by crRNA specific binding to complementary ssRNA target sequences, triggering non-specific collateral cleavage of RNA reporters. Cleavage of quenched fluorophore-labelled ssRNA reporters is detectable by measuring fluorescence with a plate reader or qualitatively under a blue-light trans-illuminator (eg. UV light). Lateral flow-based detection can be read from strips with a coloured positive/negative band using a FAM/Biotin ssRNA reporter that conjugates to anti-FAM gold nanoparticles and accumulates at the control or test lines depending on whether the reporter is intact.

### RPA primer selection and assay optimisation

Two *S. japonicum* target genes, *cox1* (cytochrome C oxidase subunit I) and *sap4* (saposin 4), were incorporated into the design of the SHERLOCK assay. *Cox1* is a mitochondrial gene and has been broadly employed as a target for accurate PCR-based detection of *S. japonicum* using stool samples [28]. The *sap4* gene is highly expressed in schistosomula and adult worms of *S. japonicum*, but not in eggs, and saposin 4 has proven to be an excellent serological diagnostic marker for detectingan early infection of *S. japonicum* [29, 30]. Two pairs of RPA primers were designed for each gene, and sensitivity of each primer pair was determined by RPA using DNA extracted from *S. japonicum* adult worms as a template. The RPA amplicons were visualised on a 2% (w/v) agarose gel (Fig 2a). A strong band at the expected size (∼125bp) was amplified by RPA using *sap4*-F1R1, *sap4*-F2R2 primers (Fig 2a, left panel) and *cox1*-F1R1 (Fig 2a, left panel), *cox1*-F2R2 (Fig 2a, right panel) primers using adult worm DNA as template. It is important to note that all the RPA negative controls appeared to show some smearing/banding in the agarose gels which could be considered as false positive reactions. Later, it was determined that the negative controls did not produce false positive results in the SHERLOCK Cas13a-detection assays. However, a strong smearing was observed in the RPA amplicon using *cox1*-F2R2 primer pair in the negative sample (H2O as template) possibly due to primer dimer (Fig 2a). Thereby the three primer pairs (*cox1*-F1R1 and *sap4*-F1R1, *sap4*-F2R2) were selected for further qPCR analysis (Fig 2b).

**Fig 2.**
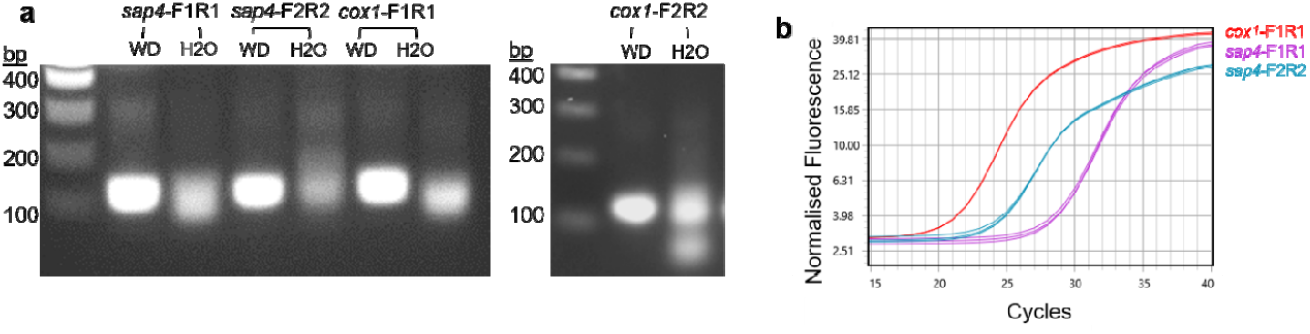
Optimisation of RPA primers for SHERLOCK detection of *S. japonicum* DNA. (a) Agarose gel images of RPA amplicons targeting genes saposin 4 (*sap*4) and cytochrome c oxidase subunit 1 (*cox1*), using the primer pairs *sap4*-F1R1, *sap4*-F2R2 (left panel) and *cox1*-F1R1 (left panel), *cox1*-F2R2 (right panel). DNA (1μl of 200ng/μl) extracted from adult *S. japonicum* worms (Philippines strain) was used as template (WD, worm DNA), H_2_O was used a negative control template; (b) qPCR amplification plot using *S. japonicum* adult worm DNA (2μl of 50ng/μl) as template to compare the detection sensitivity of the *sap4* and *cox1* genes.

To select the optimal target sequence with high amplification efficiency, we undertook qPCR assays using the *cox1*-F1R1, *sap4*-F1R1 and *sap4*-F2R2 qPCR primer pairs (Fig 2b and S1 Fig), using *S. japonicum* adult worm DNA (2μl, 50ng/μl) as the template. The *sap4*-F2R2 primers were more sensitive and effective than the *sap4*-F1R1 primers for the amplification of *sap4* (Fig 2b and S1 Fig). However, the strongest signal was observed from the reaction amplified by the *cox1*-F1R1 primer pair, indicating a higher copy number of the *cox1* gene than *sap4*, and representing a more sensitive target for the DNA detection of adult *S. japonicum*. Therefore, the *cox1*-F1R1 primer pair was employed in the subsequent development of the SHERLOCK system.

### Comparative sensitivity of the SHERLOCK system with qPCR

SHERLOCK and qPCR assays targeting *cox1* were performed using serially diluted *S. japonicum* egg DNA (20pg/μl-0.02pg/μl) as template (Fig 3). The results obtained by qPCR (Fig 3a), the SHERLOCK readout via lateral flow strip (Fig 3b), and via BioTek fluorescence plate reader (Fig 3c) all indicated an identical level of sensitivity. All three methods were capable of detecting *S. japonicum* egg DNA at a sensitivity of 0.2pg/μl. Fluorescence-based SHERLOCK detection tests normally use a fluorescence plate reader to detect the fluorescence signal, providing real-time signal kinetics. To make the fluorescence readout more field-friendly, we demonstrated that the fluorescence-based SHERLOCK assay readout could also be visualised clearly under UV light (Fig 3d).

**Fig 3.**
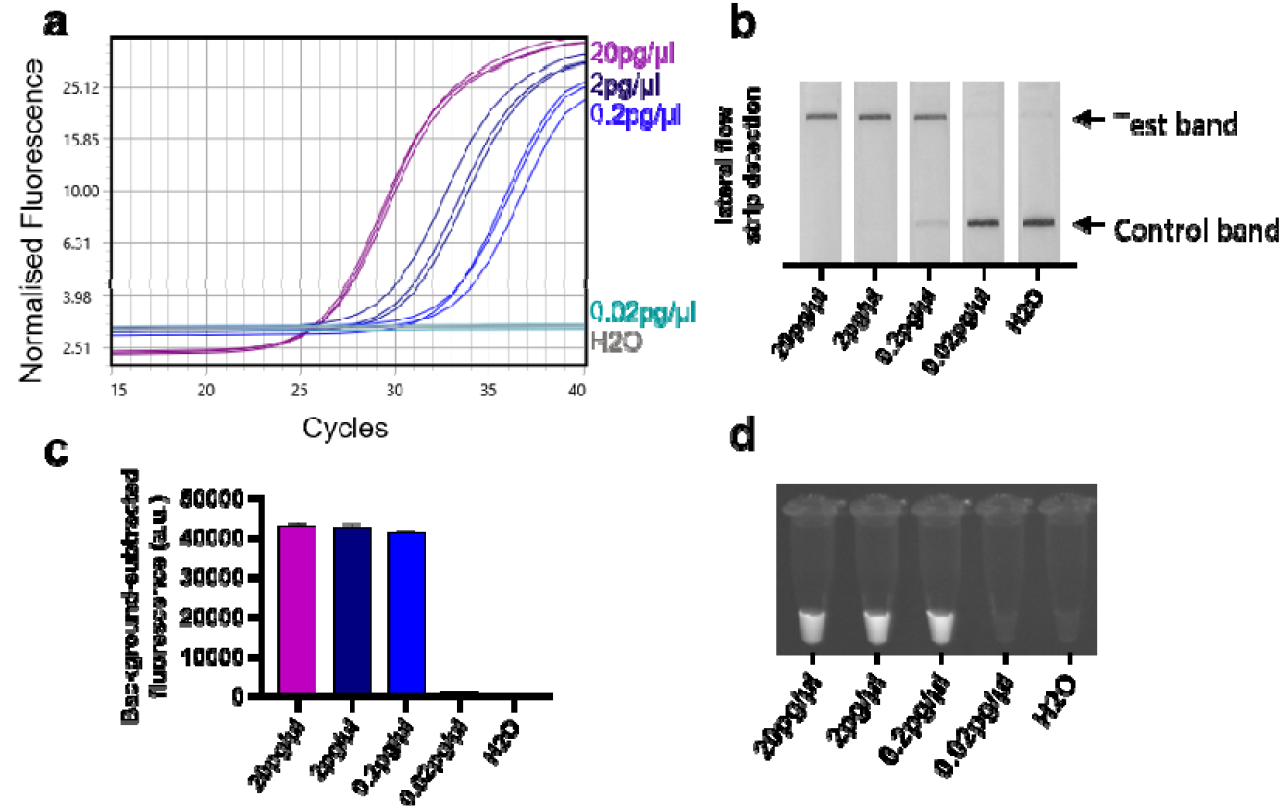
Diagnostic sensitivity of qPCR and SHERLOCK assays determined using serially diluted *S. japonicum* egg DNA samples (20pg/μl −0.02pg/μl) targeting the *cox1* gene. (a) qPCR amplification plots; (b) Colorimetric SHERLOCK assay with lateral flow readout; Fluorescence-based SHERLOCK assay with signal detected using (c) the BioTek fluorescence plate reader (mean ± SEM of 3 technical replicates), and (d) UV light.

### SHERLOCK assay validation using mouse faecal samples

To validate the accuracy and efficacy of the SHERLOCK assay, we tested DNA isolated from faecal samples obtained from individual Swiss mice (10 mice/group) infected with either a low (30/mouse) or high (70/mouse) dose of *S. japonicum* (Philippines strain) cercariae at 1-7 weeks post-infection. All the DNA samples extracted from individual mouse faeces were tested using qPCR (weeks 1-7) and SHERLOCK detection assays (weeks 4-7) targeting *cox1*-F1R1. Worms obtained from individual mice by portal perfusion at 7 weeks were counted and faecal egg number of each animal was counted at 5-7 weeks after infection (shown in S3 Table). *S. japonicum* infection was not positively identified by either qPCR or SHERLOCK at weeks 1-4 post challenge in either the low-dose or high-dose groups. However, in mice infected with the higher cercarial challenge, 7/10 mice were diagnosed as *S. japonicum*-positive at week 5 post-infection both by qPCR and the fluorescence-based SHERLOCK assay (Fig 4a). In mice that received the lower cercarial challenge dose, infection was only identified in 1/10 animals by SHERLOCK and 2/10 mice by qPCR (Fig 4b). These outcomes are consistent with the fact that sexually mature female worms of *S. japonicum* commence egg-laying after pairing with mature males around week 4 post-infection [31], with oviposition occurring between 4-6 weeks post-infection [32], resulting in egg DNA being detectable around this time. In both the low-dose and high-dose groups, *S. japonicum* infection was positively diagnosed at week 6 and week 7 post-infection in all (20/20) of the individual mice, both by qPCR and the SHERLOCK assays (via either fluorescence-based or lateral-based detection). Overall, diagnosis with the SHERLOCK assays matched well with the qPCR, results with 100% and 97.5% consensus obtained, respectively, using faecal DNA samples obtained from mice challenge infected with 70 and 30 cercariae.

**Fig 4.**
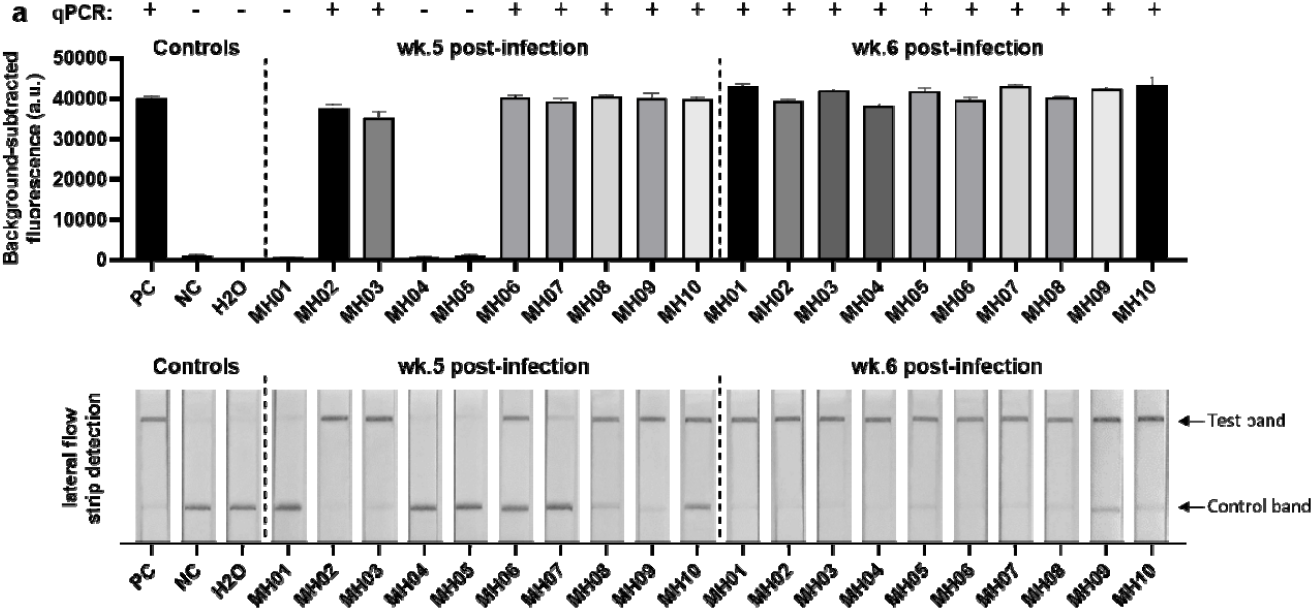
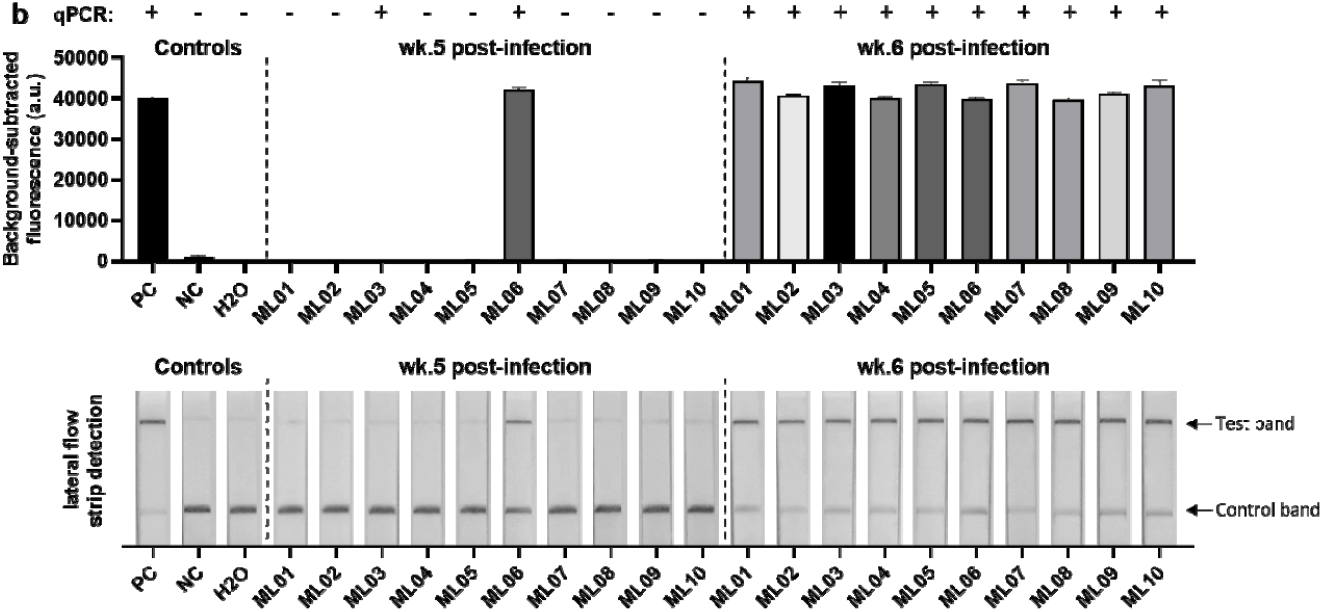
Fluorescence-based (upper panel) and lateral flow-based (bottom panel) SHERLOCK detection of *S. japonicum* (targeting the *cox1* gene) in DNA samples extracted from stools of individual mice. Mice were infected with (a) 70 *S. japonicum* cercariae and (b) 30 *S. japonicum* cercariae (Philippines strain) per mouse. Stools were collected at week 5 (wk.5) and week 6 (wk.6) post-infection. *S. japonicum* DNA (2pg/μl) extracted from eggs (Philippine strain) was used as a positive control (PC); uninfected mouse stool (negative control, NC) and H_2_O were used as negative controls. QPCR test result of each sample was shown on the top panel (+, infected; -, uninfected). Background-subtracted fluorescence values are given as the mean ± SEM of 3 technical replicates.

### SHERLOCK assay validation using clinical faecal and serum samples

To further validate the clinical diagnostic capabilities of the SHERLOCK assay, samples (sera and stool samples) collected from human subjects living in an *S. japonicum* endemic region in Northern Samar, the Philippines, were used for testing by qPCR (using the *cox1*-F1R1 primers) and the SHERLOCK assay (using *cox1*-F1R1 primers/crRNA). A detailed summary of results is presented in S4 Table. We found the SHERLOCK assay successfully detected *S. japonicum* DNA in all of a panel of both 30 qPCR-positive and 8 qPCR-negative human faecal DNA samples with 100% sensitivity and 100% specificity using both fluorescence and lateral flow readout methods (Fig 5a). Indeed, two samples that were identified as negative for *S. japonicum* infection by the Kato-Katz procedure (i.e. 0 EPG) tested positive in both the qPCR and SHERLOCK assays, emphasising the high sensitivity of the qPCR and SHERLOCK assays when testing stool samples.

**Fig 5.**
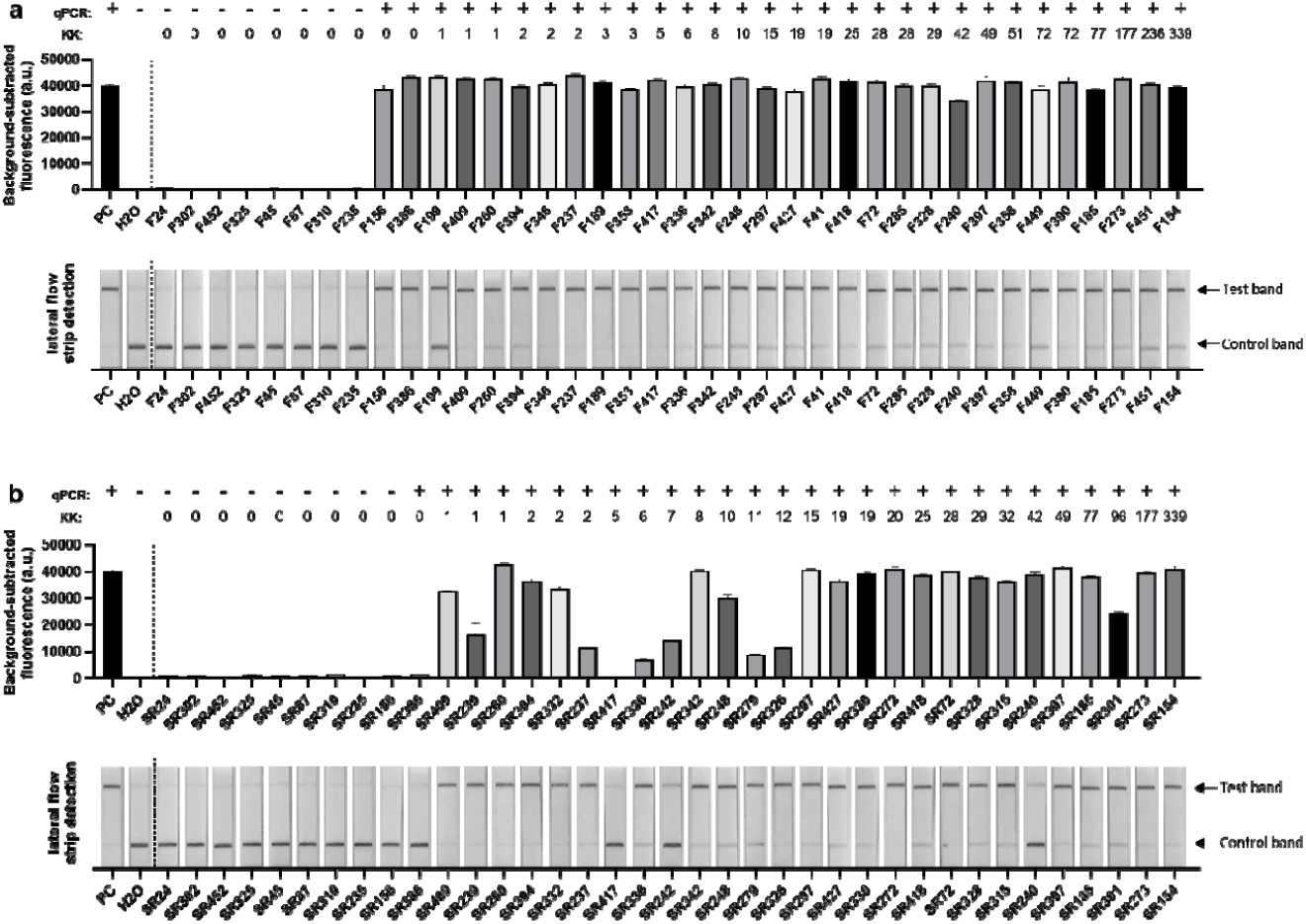
Fluorescence-based (upper panel) and lateral flow-based (bottom panel) SHERLOCK detection of *S. japonicum* DNA (targeting the *cox1* gene) in human samples. (a) faecal and (b) serum samples collected from 18 barangays endemic for *S. japonicum* infection in Laoang and Palapag municipalities, Northern Samar, the Philippines. *S. japonicum* DNA (2pg/μl) extracted from eggs (Philippines strain) and H_2_O were used as positive control (PC) and negative control, respectively. Background-subtracted fluorescence values are given as the mean ± SEM of 3 technical replicates. QPCR test result of each sample was shown on the top panel (+, infected; -, uninfected). Faecal egg counts obtained using the stool-based Kato-Katz (KK) procedure are presented as eggs per gram of faeces (EPG).

We also assessed the capacity of SHERLOCK (using the *cox1*-F1R1 primers/crRNA) to detect *S. japonicum* DNA in human serum using a panel of 28 qPCR-positive and 9 qPCR-negative serum samples (also using the *cox1*-F1R1 primers) (Fig 5b). We found 26/28 qPCR-positive serum samples also tested positive using the fluorescence-based SHERLOCK (∼93% sensitivity compared with the qPCR). However, the mean background-subtracted fluorescence values of the ‘positive’ serum samples (with a fluorescence value <50% of the positive control) was variable compared with the results obtained with the human and mouse stool samples. In those samples with a lower than expected mean fluorescence value, it is probable that RPA pre-amplification did not reach the saturation point and thus Cas13 detection proceeded at a slower rate, likely due to very low target copy numbers present in the samples. Nevertheless, samples with low mean fluorescence were still considered ‘positive’ as they provided a fluorescence readout that was significantly higher than the negative controls (**** *P*<0.0001, one way ANOVA).

It is notable that of the 28 qPCR-positive sera samples, 26 clearly tested positive by the fluorescence-based SHERLOCK assay. However, 2 of the 26 samples presented a faint test line in the lateral-based detection assay; although rare, this could be considered as an ambiguous or open to interpretation result; this was also recorded with one mouse DNA sample (MH07; at 5 weeks post-infection) (Fig 4a). This suggests that the lateral flow detection may have slightly reduced sensitivity compared with the fluorescence-based SHERLOCK detection, which can provide a clearer and more easily interpretable result. The SHERLOCK assay had 100% specificity compared with the qPCR using the serum samples as the 9 qPCR-negative sera were also negative tested using both the fluorescence and the lateral flow-based detection methods.

In an effort to identify an improved target gene for testing the serum samples, we used an alternative fluorescence-based SHERLOCK assay, incorporating *sap4*-F2R2 RPA primers/crRNA, to test the same panel of human sera from the Philippines (S2 Fig). However, a positive diagnosis was only obtained with 3 of the 28 qPCR-positive (using *cox1*-F1R1 primers) serum samples using the *sap4* SHERLOCK assay, demonstrating its very low sensitivity (∼11%) compared with the qPCR. Clearly, the SHERLOCK assay targeting *cox1*-F1R1 provided a considerably higher level of diagnostic sensitivity and accuracy than the *sap4*-F2R2 primer pair achieved.

## Discussion

CRISPR-Cas13 technology represents a powerful new generation diagnostics platform that had not been applied for the detection of *Schistosoma* spp. infections or, indeed, any other helminthic disease of humans or animals. We demonstrate here, for the first time, the capacity of a SHERLOCK assay to diagnose an infection caused by *S. japonicum*. The diagnostic assay achieved robust sensitivity when applied to stool samples obtained from mice experimentally infected with a high and low dose challenge of *S. japonicum* cercariae, and human clinical samples (stool and sera) collected from schistosomiasis-endemic areas (Laoang and Palapag municipalities, Northern Samar), in the Philippines. The results we present here reinforces the potential of SHERLOCK and the CRISPR diagnostic platform to fulfill the urgent requirements for new diagnostic tools to better assess and assist in the control and elimination of schistosomiasis and other parasitic NTDs globally.

The most commonly utilised readout methods of SHERLOCK are fluorescence-based detection via a plate reader and colorimetric detection via a lateral flow strip; we tested total 75 human samples and 80 animal samples using both methods to assess the merits of each. Fluorescence-based detection using a plate reader is more suitable for mass-screening as the interpretation of positive and negative results can be clearly defined; however the requirement of a fluorescence plate reader means this method is less field-friendly, especially for use in low resource, rural environments. To make the florescence-based SHERLOCK readout more user-friendly, we showed that the detection signal was also visible by the naked eye in-tube under UV light, and provided the same level of sensitivity as the fluorescence plate reader (Fig 3). To reduce user bias during the detection process, a potential solution could be to incorporate a smartphone app that can aid in the interpretation of results [21], a UV reader device that can be added to a standard smartphone [33], which are all feasible to use. Lateral flow-based detection methods have the benefit of being portable and easy to use. For the most part, the interpretation of results is straight-forward, but we found that some of the analysed samples occasionally produced a very faint test line when we analysed samples with very low *S. japonicum* infection intensities. Such anomalies may cause difficulties in interpreting these as definitively positive, an observation that has also been noted in another study [34]. Quantification of band intensities using image analysis software may allow for more clearly defined cut-off point to be set, but could complicate the analysis.

In developing the SHERLOCK assay for the detection of *S. japonicum*, we considered two key target genes, *cox1* and *sap4. Cox1* is a haploid-inherited mitochondrial gene that is very concertedly and repetitively represented in schistosomes [28], and has been employed to identify and differentiate infections among *S. mansoni, S. haematobium, S. japonicum, S. mekongi* and *S. bovis* [35, 36]. Future development of the SHERLOCK assay based on species-specific *cox1* could potentially extend the platform for the accurate diagnosis of other schistosome species and other helminths. We show here that *cox1* is also a robust diagnostic target for *S. japonicum* identification using SHERLOCK, resulting in 100% and 93% sensitivity, respectively, targeting clinical stool and serum samples compared with qPCR.

The *sap4* gene encodes the saposin-like protein SAP4, a molecule suspected to be critical in the lysis of ingested red cells in the schistosome gut, playing an important role in regulating host immune response [37]. SAP4 has been shown to be an excellent target for serological diagnosis of schistosomiasis using ELISA, capable of identifying infected individuals in cohorts from Asian schistosomiasis-endemic areas, both in China and the Philippines [29, 30]. We assessed the value of *sap4* as a target gene in our SHERLOCK assay by testing the 28 human serum samples that tested positive by qPCR with *cox1* primers, but only 3/28 (∼11% sensitivity compared with the qPCR) tested positive for *S. japonicum* using the *sap4-*based assay (S2 Fig). This poor level of sensitivity may reflect a low transcript level of *sap4* present in patient sera. SAP4 has proven an excellent serological diagnostic marker, being consistently released in worm vomitus into host blood [38], thereby inducing a strong host immune response resulting in high levels of anti-SAP4 antibody which is readily detectable by ELISA. However, the *sap4* gene may not be a good target for a nucleic acid based diagnostic assay, given that gene abundance levels are frequently not reflected in protein abundance levels, and vice versa [39]. In light of a previous study of ours results showing that *sap4* is not transcribed in *S. japonicum* eggs [30], we did not test the *sap4* SHERLOCK assay using stool samples.

As this is the first application of CRISPR-Cas13 based diagnostics to detect a parasitic helminth infection, we acknowledge the limitations of our assay design and the improvements that will be required before it can be applied as a clinical POC application in resource-limited settings. Based on the CRISPR-based diagnostics literature and our study results, we have identified several strategies to develop our assay towards the goal of its field-deployment and testing its diagnostic effectiveness in the future:

1. Further improvements to the sensitivity of the SHERLOCK assay, particularly during the early stages of an infection, could be made by testing additional gene targets such as the mitochondrial nad1 gene and the retrotransposon, SjR2 [40] and applying a wider panel of crRNA/RPA primers.
2. The SHERLOCK assay procedure could be simplified by incorporating a rapid DNA extraction method or by combining the RPA reaction with Cas13 detection (Cas13 protein, crRNA and reporter) in one single reaction to shorten the processing time. This would reduce the equipment, resource costs and the technical requirements, thereby making the assay faster and more field-friendly. In the our protocol, we used an extended incubation time for the RPA and CRISPR-Cas13 detection, but these could be reduced to as little as 5 minutes for each step as described [41]. Combining the RPA pre-amplification and CRISPR-Cas13 detection into one single reaction (“one-pot” detection) could also reduce the reaction time, as well as reducing the risk of introducing contaminants. One-pot CRISPR-Cas detection has been successfully implemented for the detection of the Zika and Dengue viruses [42] as well as malaria [22]. However, protocol optimisation can be difficult and reaction viscosity may reduce sensitivity [41]. The requirement for effective DNA extraction is critical for nucleic-acid based detection methods; compared with PCR, RPA and LAMP amplification methods that are used in CRISPR-Cas detection have been shown to have increased tolerance to inhibitors present in urine, serum and plasma [43, 44]. One method that shows promise uses DNA dipstick technology incorporating wax-impregnated Whatman No. 1 filters to extract DNA from human clinical samples in 30 seconds without the need for electrical equipment or pipettes [45]. We have successfully applied the DNA dipstick in qPCR and LAMP assays on adult *S. japonicum* and eggs, and schistosome-infected snails [46]. The manufacture of dipsticks uses low cost, easily accessible materials, and the speed and ease-of-use of the extraction process makes this method especially suitable for use in low-resource settings [45]. Other reports have combined rapid DNA extraction procedures with CRISPR-Cas detection using heat and/or chemical additives to inactivate nucleases and inhibitors [19, 22]. Further research in this developing area will be required to expedite our SHERLOCK assay without sacrificing sensitivity.
3. Protocol changes, such as using LAMP-based CRISPR-Cas12 mediated detection, could be considered. Both RPA and LAMP are commonly used in CRISPR-Cas detection protocols as pre-amplification of the target gene significantly enhances sensitivity [47]. RPA is often chosen because of its speed and the low amplification temperature required which facilitates one-pot detection. Although LAMP primer design is more complex, it exhibits greater specificity [48], and there is some evidence that LAMP may have greater sensitivity than RPA and have a greater tolerance of inhibitory substances [49]. LAMP is more commonly coupled with CRISPR-Cas12 mediated detection, and one-pot detection coupling LAMP and Cas12b is possible [50].

At present, diagnosis of schistosomiasis is still often based on microscopic analysis of stool and urine samples using the Kato Katz procedure, which though relatively simple and inexpensive, lacks sensitivity, particularly when infection prevalence/intensity levels are low [51]. Relying on low sensitive diagnostics has proven to lead to underestimation of infection rates [52] which has serious implications for disease management. Nucleic acid detection methods such as PCR have been developed for schistosomiasis and frequently achieve a much higher degree of sensitivity [28] but are rarely used for clinical diagnosis within endemic countries due to their requirements for expensive laboratory equipment and trained personnel. Here we demonstrate that using the CRISPR-Cas13 SHERLOCK platform we were able to detect a schistosome infection with a level of sensitivity similar to qPCR-based assays (93-100%) and able to detect as little as 0.2 pg/μl *S. japonicum* egg DNA (Fig 3). When considering its ease of access, our SHERLOCK assay has many benefits over qPCR. SHERLOCK can be performed at a constant temperature of 37°C, which is easily achievable using a simple, portable heat block and bypasses the requirement of a thermal cycler. Indeed, no specialized equipment is required and, with multiple field-friendly readout methods available including lateral-flow or in-tube fluorescence detection by UV light, the interpretation of results is easy and straight-forward. The field of CRISPR-Cas based diagnostics is growing rapidly and collective research efforts have already led to many improvements in assay design including one-pot detection, DNA extraction from crude samples, and increased accuracy at the POC and in the field [47]. The technology is readily adaptable, and capable of incorporating multiplex assays that can distinguish between individual species, specific to 1-2 single nucleotide polymorphisms (SNPs) [21]. As development continues, CRISPR based diagnostics show great promise and move closer to POC application in resource-limited settings.

The development of a POC test that has comparable sensitivity to qPCR would be an invaluable tool in the global fight against schistosomiasis. We demonstrate here a SHERLOCK assay to detect *S. japonicum*, but the technology should be readily adaptable to detect other schistosome and helminth species. The procedure has considerable potential to be developed into a POC tool for the early detection of schistosomiasis, offering a higher level of sensitivity than any POC test in use today. Schistosomiasis and other neglected helminthic diseases cause severe disability contribute to malnutrition, hinder growth and negatively impact on the outcomes of pregnancy, on physical performance, productivity and cognitive development, and are often fatal; children are affected disproportionately. Accurate and rapid CRISPR-based POC diagnostics are crucial for immediate and effective treatment and for the prevention of any severe disease. Access to sensitive POC diagnostics will provide accurate data on schistosomiasis status and tracking that can feed into essential risk mapping for quick responses in areas of increased transmission to better manage and respond to disease outbreaks. Rapid and early POC detection of cases is critically important if the ultimate goal of schistosomiasis elimination is to be achieved. CRISPR-based diagnostics show great potential as a new generation of POC diagnostic tools that can be optimised for the detection of a range of NTDs. Access to these resources will result in important diagnostic outcomes of major and timely public health relevance, particularly as recent global disruptions due to COVID-19 have resulted in the interruption of control programs for schistosomiasis and other NTDs [53] worldwide.

The SHERLOCK assay we developed in this study is capable of detecting a *S. japonicum* infection at a similar level of sensitivity and specificity comparable to qPCR, using both faecal and serum clinical samples. With its versatility, sensitivity, and its potential to be an inexpensive, portable POC tool, SHERLOCK can become a major and dominant player as a next-generation diagnostic platform for helminthic infections. It will provide the ammunition to further extend this sensitive, field-friendly diagnostics approach for future wide-scale application to other parasitic NTDs globally. The CRISPR-Cas12/13 based diagnostics platform we are pioneering fits well with the unmet and urgent requirements outlined by the WHO [4] for rapid mapping of helminthic diseases, monitoring helminth control programs and assessing elimination targets. As highlighted by the new roadmap for NTDs 2021-2030 released by the WHO [3], POC testing enables control programs to target only infected subjects for drug treatment which can effectively reduce drug costs, increase acceptance of treatment, and decrease the risk of drug resistance. Future investigations are needed to optimise performance of SHERLOCK in field settings but accelerated research will open new opportunities for CRISPR diagnostics in clinical, surveillance and research applications. The novel *S. japonicum* SHERLOCK assay presented in this study emphasises the promise of CRISPR diagnostics and demonstrates the vital role it may play in the global fight against schistosomiasis and other NTDs.

## Methods

### Ethics statement

All experiments undertaken in this study were approved by the Animal and Human Ethics Committee (ethics number P3706) of the QIMR Berghofer Medical Research Institute and the Ethics Committee of the Research Institute for Tropical Medicine (RITM), Manila, the Philippines. The study was performed according to the guidelines of the National Health and Medical Research Council of Australia, which were published in the Australian Code of Practice for the Care and Use of Animals for Scientific Purposes, 7th edition, 2004 (www.nhmrc.gov.au). All work related to live *S. japonicum* life cycle stages was conducted in quarantine-approved facilities.

### Mouse infection and faecal sample collection

Six-week-old female ARC Swiss mice (10 mice/group) were infected percutaneously with either a low dosage (30/mouse) or high dosage (70/mouse) of *S. japonicum* (Philippines strain) cercariae. Faeces were collected from individual mice weekly 1-7 weeks post infection for DNA extraction. Egg counting of individual faecal samples was undertaken at 4-7 weeks post infection as described [54]. Mice were sacrificed at 7 weeks post infection and adult worms obtained by portal perfusion were counted. Uninfected mice were used as negative controls.

### Clinical sample (faeces and serum) collection

Clinical samples (faeces and blood) were collected from subjects residing in 18 barangays (villages) endemic for schistosomiasis japonica in Laoang and Palapag municipalities, Northern Samar, the Philippines, as described [55]. Briefly, individual serum samples were obtained by centrifuging blood samples in serum separation tubes at 1500g for 10 min after prior incubation at room temperature for 30 min. Age and gender information for each participant were recorded at the time of sampling. Individual stool (fixed in 80% ethanol) and serum samples, collected from each subject, were shipped with dry ice to QIMRB, Australia for DNA extraction and further analysis. The Kato-Katz test was performed on all faecal samples as described [56] to estimate infection prevalence and infection intensity was presented as the number of eggs per gram of faeces (EPG).

### DNA extraction from mouse and human samples

Genomic DNA was isolated from mouse faecal samples using the QIAamp DNA Mini kit (Qiagen, Hilden, Germany). Stool (200mg/sample) was first washed with ddH_2_O, then 500μl ROSE buffer was added and samples were manually homogenized using a toothpick as described [55]. Homogenates were incubated at 95°C for 10 min and centrifuged at 4000g for 5 min. 200μl of the resulting supernatant was transferred into a new tube along with 25μl of proteinase K, and the remaining DNA isolation steps were carried out following the Qiagen protocol.

Genomic DNA was extracted from the ethanol-fixed human faecal samples using the Maxwell®16 Instrument (Promega, Wisconsin) and the Maxwell®16 LEV Plant DNA kit as previously described [55]. Human serum DNA was extracted from 2mL of each serum sample using the ChemagicTM360 instrument (PerkinElmer Inc., Massachusetts).

### RPA and crRNA primer design

RPA primers were designed targeting recognised diagnostic genes for schistosomiasis including the mitochondrial *cox1* (cytochrome C oxidase subunit 1) [28] and *sap4* (coding saposin-like protein 4) [57]. The NCBI Primer Basic Local Alignment Search Tool (BLAST) was used for RPA primer design according to the following criteria: 1) 30-35 nucleotides in length; and 2) amplicon length of ∼125bp targeting a ∼28bp crRNA binding site. A T7 promoter sequence (GAAATTAATACGACTCACTATAGGG) was incorporated into the 5’ end of the forward primer to enable in vitro transcription by T7 polymerase during the SHERLOCK detection [16]. Primers were synthesized by Integrated DNA Technologies (IDT, Singapore).

After RPA primer optimisation, crRNAs were designed targeting *cox1* and *sap4*, synthesised by IDT, resuspended in nuclease-free water and stored at −20°C. The full list of oligonucleotides (including the RPA primers, qPCR primers, crRNA) used in this study is provided in S1 Table.

### Recombinase polymerase amplification (RPA)

RPA reactions were performed using the TwistAmp Basic (TwistDx, Maidenhead, UK) kit according to the manufacturer’s instructions, with the following modifications. Magnesium acetate was added prior to sample input, and each 50μl TwistAmp reaction was divided into 5x 10μl reactions, each containing 9μl of the reconstituted RPA reaction and 1μl DNA sample. Reactions were incubated at 37°C in a thermal cycler for 1 h. After 7 min initial incubation, samples were removed, vortexed, centrifuged briefly, and then returned to the cycler for the remaining incubation time. For preliminary RPA primer screening, reactions were purified using a QIAquick PCR Purification Kit (Qiagen) and then visualised on a 2% (w/v) agarose gel.

### Detection using LwCas13a

Fluorescent detection assays were performed in triplicate in a 20μl reaction volume containing 44nM LwCas13a (GenScript, Piscataway, USA), 10ng crRNA (IDT), 125nM fluorescent Poly-U reporter (IDT, S2 Table), 1mM rNTP mix (New England Biolabs; NEB, Ipswich, USA), 20mM HEPES, 9mM MgCl_2_, 0.6μl Murine RNase inhibitor (NEB), 0.5μl T7 RNA Polymerase (LGC Biosearch Technologies, Middlesex, UK) and 1μl RPA product. Reactions were incubated at 37°C for 1h on a BioTek Synergy H4 multi-mode plate reader (Agilent, Santa Clara, USA) with kinetics measured every 5min. Background-subtracted fluorescence was calculated by subtracting the absolute fluorescence values of no-template control wells (ddH_2_O replacing the RPA product) from sample well fluorescence values. To compare samples across runs, sample background-subtracted fluorescence values were normalised relative to positive control (2ng *S. japonicum* worm cDNA) fluorescence values which were set to 40,000 a.u. Data are presented as the mean ± SE for n=3 technical replicates. Some reactions were also visualised under UV light and the results captured by smart phone camera.

Lateral flow assays were set up with the same reaction components as the fluorescent detection assays, substituting the fluorescent reporter with 250nM FAM-Biotin Poly-U reporter (IDT, S2 Table). After incubation at 37°C for 1 h, 80μl HybriDetect 1 assay buffer (Milenia Biotec, Gießen, Germany) was added to 20μl of the Cas13a reaction and run on HybriDetect 1 lateral flow strips (Milenia Biotec, Gießen, Germany). The strip was incubated at room temperature for 2-3 min and the result was recorded by camera.

### Real-time PCR

Real-time PCR (qPCR) was used as a gold standard to determine the sensitivity of the SHERLOCK system using the identical samples tested using the SHERLOCK detection assays. Sequences of the qPCR primers used in this study are shown in S1 Table. Reaction mixtures comprised: 10μl QuantiNova SYBR Green PCR Master Mix (Qiagen), 200nM each of the forward and reverse primers, 2μl template DNA, and ddH_2_O to a final reaction volume of 20μl. The following cycling conditions were used: initial denaturation at 95°C for 5 min, followed by 35 cycles of denaturation at 95°C (30 s), annealing at 59°C (30 s), and extension at 72°C (30 s). The PCR was performed using the Mic qPCR cycler and software (Bio Molecular Systems, Upper Coomera, Australia). Detection of target sequence was indicated by cycle threshold (Ct) values as previously described [58, 59]. For each run, the non-template (ddH_2_O) reaction was used as negative control and the *S. japonicum* DNA extracted from either eggs or adult worm pairs was used as positive control. A positive qPCR result was defined as successful amplification (positive Ct value) in >80% technical replicates. To compare runs, sample qPCR values were adjusted relative to the positive control (threshold set so the positive control mean Ct=25.5). Data were analysed using a dynamic baseline correction with extensive exclusion parameters and manual exclusion of samples with an abnormal melt curve.

### Statistical analysis

Statistical analyses were conducted using Prism GraphPad (Version 7, GraphPad Software, La Jolla, CA, USA). Each experiment was performed in duplicate and all data are presented as the mean ± SE of technical replicates. Comparisons between groups were performed to determine statistical significance using One-way ANOVA with Dunnett’s correction for multiple comparisons. The relative sensitivity and specificity of the SHERLOCK assays were determined using qPCR (*cox1*-F1R1 primers) as the reference test.

## Data Availability

All data produced in the present study are available upon reasonable request to the authors

## Acknowledgments

We thank Mary Duke from QIMR Berghofer Medical Research Institute for the maintaining of the *S. japonicum* life cycle and the provision of parasite materials for this study. *Oncomelania hupensis quadrasi* snails were provided by the NIAID Schistosomiasis Resource Center of the Biomedical Research Institute (Rockville, MD) through NIH-NIAID Contract HHSN272201700014I for distribution through BEI Resources. We thank all research participants and the local Philippines field and clinical staff for their kind assistance allowing collection of the clinical samples used in this study. This work received support from an Australian Infectious Disease Research Centre Seed Grant and Grants (APP1194462, APP2008433) from the National Health and Medical Research Council of Australia.

## Author Contributors

HY, HS, JDF, and DPM contributed to study design; SM and HY performed all experiments; HY, SM, HS, JDF contributed to development of methodology; CAG, PC, RMO and MKJ provided reagents, contributed to sample collection and data analysis; SM, HY, DPM, JDF and MKJ, drafted and revised the manuscript. HY and DPM are responsible for funding acquisition and project administration. All authors contributed to the article and approved the submitted version.

## Supporting Information

**S1 Fig.**
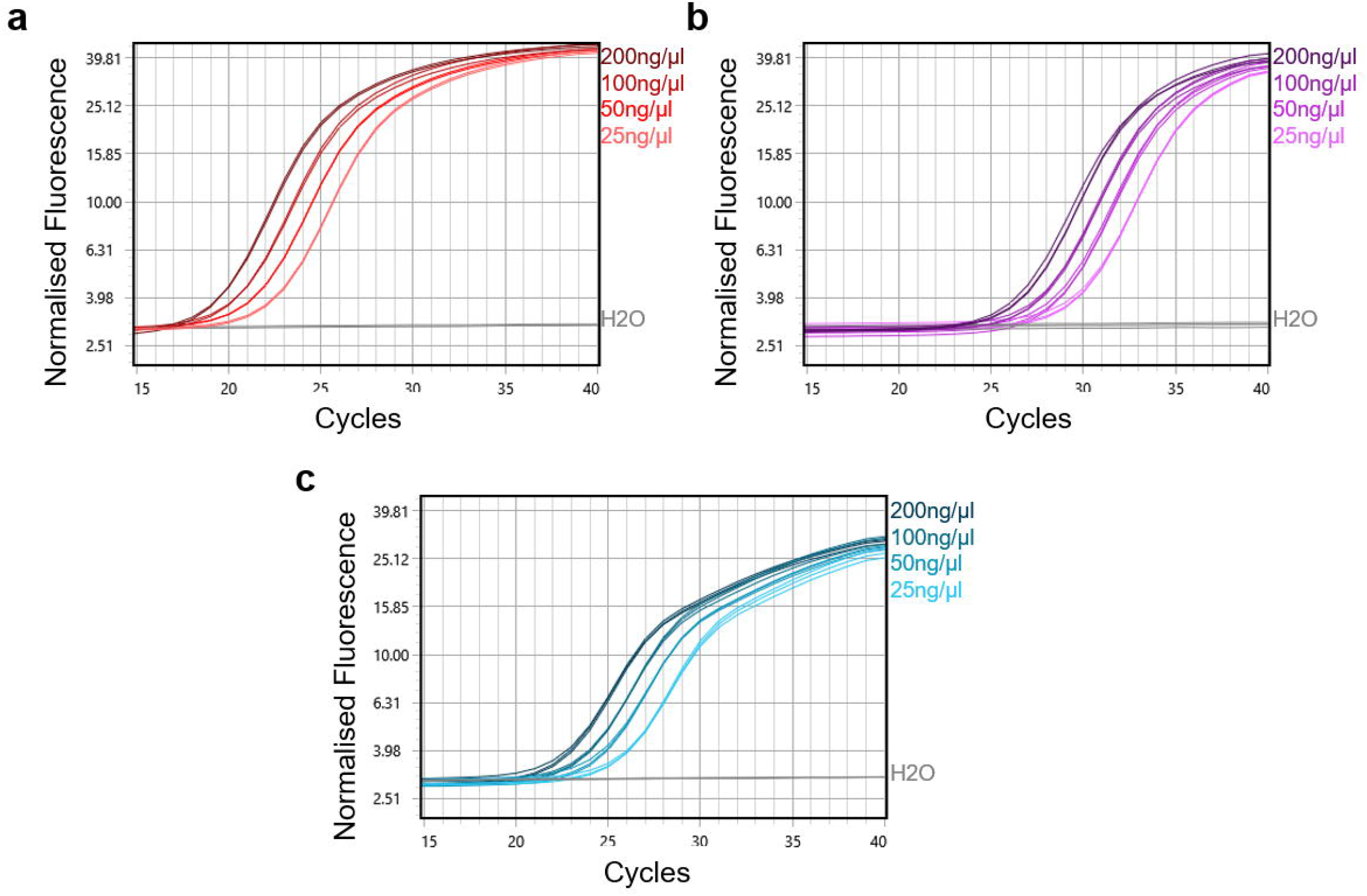
Analytical sensitivity of qPCR assays using different gene targets as determined using serially diluted *S. japonicum* adult worm DNA (200ng/μl-25ng/μl). Amplification plots are shown for primer pairs (a) *cox1*-F1R1, (b) *sap4*-F1R1, and (c) s*ap4*-F2R2.

**S2 Fig.**
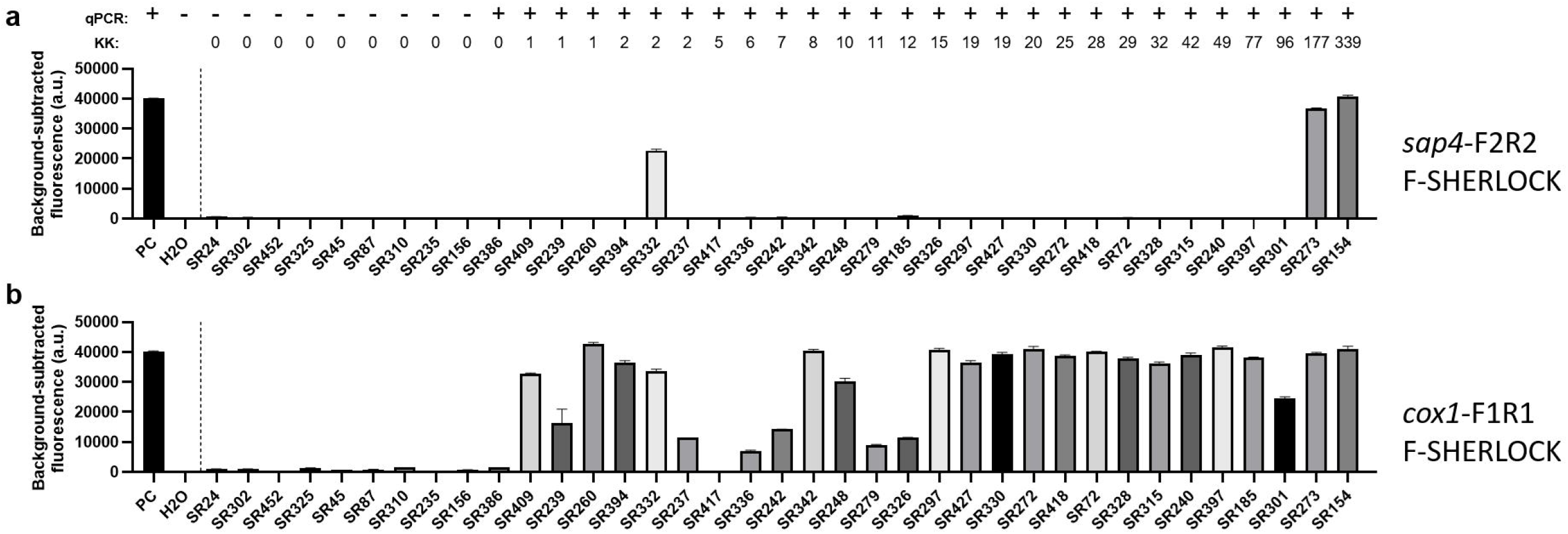
Fluorescence-based SHERLOCK detection of *S. japonicum* in human serum samples collected from 18 barangays endemic for *S. japonicum* infection in the Philippines. Targeted genes were: (a) *sap4* (using *sap4*-F2R2 primers/crRNA) and (b) *cox1* (using *cox1*-F1R1 primers/crRNA). *S. japonicum* DNA (2ng/μl) extracted from adult worms (Philippines strain) and H_2_O were used as positive control (PC) and negative control, respectively. Background-subtracted fluorescence values are given as mean ± SEM of 3 technical replicates. QPCR test result of each sample was shown on the top panel (+, infected; -, uninfected). Faecal egg counts used the stool-based Kato-Katz (KK) method and are presented as eggs per gram of faeces (EPG).

**S1 Table. Oligonucleotides used in this study**.

**S2 Table. RNA cleavage reporters used in this study**.

**S3 Table. Mouse faecal sample testing results**. Sample qPCR values normalised relative to 2ng *S. japonicum* worm DNA positive control (threshold set so Ct=27), using mic PCR Dynamic baseline correction with extensive exclusion parameters and manual exclusion of samples with abnormal melt curve. Positive (+) qPCR result defined as successful amplification (positive Ct) in >80% technical replicates. Positive (+) lateral flow (LF) SHERLOCK result defined as strong positive ‘test’ band. Positive (+) fluorescence (F) SHERLOCK result defined as mean background-subtracted fluorescence value of sample significantly different to no-template control (**** P<0·0001, one way ANOVA). NA, not applicable.

**S4 Table. Human serum and faecal sample testing results**. Sample qPCR values normalised relative to 2ng *S. japonicum* worm DNA positive control (threshold set so Ct=25.5), using mic PCR Dynamic baseline correction with extensive exclusion parameters and manual exclusion of samples with abnormal melt curve. Positive (+) qPCR result defined as successful amplification (positive Ct) in >80% technical replicates. Positive (+) lateral flow (LF) SHERLOCK result defined as strong positive ‘test’ band. Positive (+) fluorescence (F) SHERLOCK result defined as mean background-subtracted fluorescence value of sample significantly different to no-template control (**** P<0.0001, one way ANOVA). NA, not applicable. Patient IDs shown in this Table were not known to anyone outside the research group

